# Initial Observation of Contrast Profiles for 2D and 3D MRI Sequences in MR-Guided Radiation Therapy for Locally Advanced Pancreatic Cancer

**DOI:** 10.1101/2023.04.07.23288201

**Authors:** Gobind S. Gill, Brady Hunt, Rongxiao Zhang, Benjamin B. Williams, Bassem I. Zaki

## Abstract

**Purpose:** In our experience treating locally advanced pancreatic cancer (PC) with magnetic resonance-guided radiation therapy (MRgRT), the true-fast imaging with steady-state free precession (TRUFI) sequences used to generate real-time 2D MRI (2D cine) impart differing intensities for relevant structures when compared to the pre-treatment high resolution 3D MRI (3D MRI). Since these variations can confound target tracking selection, we propose that an understanding of the differing contrast profiles could improve selection of tracking structures.

**Methods and Materials:** We retrospectively reviewed both 2D cine and 3D MRI images for 20 patients with PC treated with MRgRT. At simulation, an appropriate tracking target was identified and contoured on a single 3mm sagittal slice of the 3D MRI. This sagittal slice was directly compared to the co-registered 7mm 2D cine to identify structures with notable discrepancies in signal intensity. The 3D MRI was then explored in additional planes to confirm structure identities. For quantitative verification of the clinically observed differences, the pixel intensity distributions of 2D cine and 3D MRI DICOM image datasets were statistically compared.

**Results:** In all patients reviewed, arteries (aorta, celiac, SMA, HA) appeared mildly hyperintense on both scans. However, veins (PV, SMV) appeared hyperintense on 2D cine but isointense on 3D MRI. Biliary structures appeared mildly hyperintense on 2D cine but starkly hyperintense on 3D MRI. The pixel intensity distributions extracted from 2D cine and 3D MRI images were confirmed to differ significantly (two sample Kolmogorov-Smirnov test; test statistic =0.40; p <0.001).

**Conclusions:** There are significant variations in image intensity between the immediate pre-treatment 2D cine when compared to the initial planning 3D MRI. Understanding variations of image intensity between the different MRI sequences used in MRgRT is valuable to radiation oncologists and may lead to improved target tracking and optimized treatment delivery.

## Introduction

### Neoadjuvant Therapy for Pancreatic Cancer

Pancreatic cancer (PC) is a highly lethal malignancy with a 5-year survival rate of 11%.^1^ At diagnosis, nearly 50% of patients present with locally advanced pancreatic cancer (LAPC). For PC as a whole, an R0 resection seems to be the greatest positive factor affecting outcomes. While systemic treatment of PC has improved, surgical resection rates remain low, and patients with borderline resectable or unresectable LAPC still have poor outcomes.

It is hypothesized that neoadjuvant treatment can improve rates of R0 resection, local control and survival^2^; however, there have been conflicting results regarding the optimal neoadjuvant regimen. A phase 3 trial failed to demonstrate a benefit with the addition of neoadjuvant conventionally fractionated RT (CFRT) to chemotherapy following induction chemotherapy.^3^ However, a propensity-matched retrospective review comparing neoadjuvant stereotactic body radiation therapy (SBRT) to CFRT demonstrated an improved overall survival with SBRT.^4^ Other studies have demonstrated higher biological effective dose (BED) with SBRT as a predictor of improved overall survival (OS).^5^ A meta-analysis of over 1000 patients across 19 trials concluded that SBRT did not have superior outcomes compared with conventional RT, but they postulated that dose escalation would likely have a clinical benefit.^6^ This echoes the sentiment of other studies regarding the dosimetric feasibility of dose-escalating CT-based SBRT for LAPC on linacs.^7^ Unfortunately, the success of neoadjuvant CT-based SBRT for LAPC has been limited due to excess dose resulting in toxicity to the OARs and use of non-ablative doses that did not clearly demonstrate improved OS for inoperable PC.^8,9^

### MR-Guided Adaptive SBRT

MR-guided RT (MRgRT) is a novel approach utilizing MRI scans, acquired both prior to and continuously during treatment to enable soft tissue patient positioning, beam gating based on soft tissue tracking, and online adaptive planning. Stereotactic magnetic resonance-guided adaptive radiation therapy (SMART) utilizes the unique aspects of MRgRT mentioned above to push radiation target doses higher while respecting established OAR constraints.

Preliminary data of SMART for LAPC from a single institution demonstrated grade 3 toxicities at only a ∼3% rate with a median follow up of 10.3 months.^10^ A multicenter phase II trial, which recently closed to accrual, implemented SMART in an effort to increase BED to 100 Gy, using an integrated 0.35T MR-cobalt or MR-linac system (ViewRay Inc., MRIdian®, Denver, CO, USA).^11^ Details regarding the MR-cobalt and MR-linac technologies and associated workflow have previously been reported.^12,13^ The GTV was defined as the tumor. The CTV was defined as GTV with or without regional lymph nodes and a 3 mm margin was used for PTV construction. The prescription dose was 50.0 Gy in 5 fractions.

Per protocol, prior to every treatment, a volumetric MRI scan (3D MRI) was performed to visualize anatomic changes and predict radiation dose to both target volumes and OARs as per the original plan. When OAR constraints were violated because of anatomic changes, the study required that on-table adaptive planning be performed to ensure OAR constraints were met; optimizing target volume coverage was a secondary objective. The original plan was not delivered if: 1) a GI OAR constraint was violated 2) coverage of the CTV was <85% by the 47.5 Gy isodose line 3) there was a favorable shift in CTV and OAR doses in the adaptive plan such that the CTV coverage by the 47.5 Gy isodose line is improved by 10% or more compared with the original plan. On-table plan adaptation, plan evaluation, and adaptive quality assurance occurred while the patient remained in the treatment position. During treatment delivery, sagittal planar MR images (2D cine) were continuously acquired during treatment delivery at either 4 or 8 frames per second and the radiation beam was automatically paused when the tracked pancreatic tumor moved, typically due to respiratory motion, out of a defined gating boundary.

### MR Sequences

True fast imaging with steady-state free precession (TRUFI) is a T1/T2 weighted MRI sequence which utilizes steady states of magnetizations through gradient reversal echoes and short repetition time between successive excitation pulses thereby offering high signal-to-noise ratio per unit time.^14^ Work has been done for decades to improve these methods and preserve high signal-to-noise in the presence of motion.^15^ An application in radiation therapy of TRUFI is “real time” tracking that is employed in MRgRT by observing motion on a 2D sagittal slice.

Due to the different sequences to visualize the 2D real time cine and the initial pretreatment 3D setup MRI, there is a large discrepancy in the intensity of these structures. Based on this background, we aimed at devising a qualitative guide to help establish a rule of thumb for physicians using the MRgRT system.

## Methods and Materials

### Accessing Image Sets

With IRB approval, DICOM files for both 2D cine images and 3D MRI images stored in the treatment planning system (TPS) were retrospectively accessed for patients with PC (n=20) treated on the MR-linac. At the time of simulation, an appropriate tracking target was identified and contoured on a single 3mm sagittal slice of the 3D MRI. At our institution, the default structure selected for tracking is often the visible tumor.

This sagittal slice from the 3D MRI was then directly compared to the co-registered 7mm 2D cine to identify structures with notable discrepancies in signal intensity paying particular interest to arteries, veins, and biliary structures. The 3D MRI was then explored in additional planes to confirm structure identities. This process is summarized in Figure 1.

**Figure 1.**
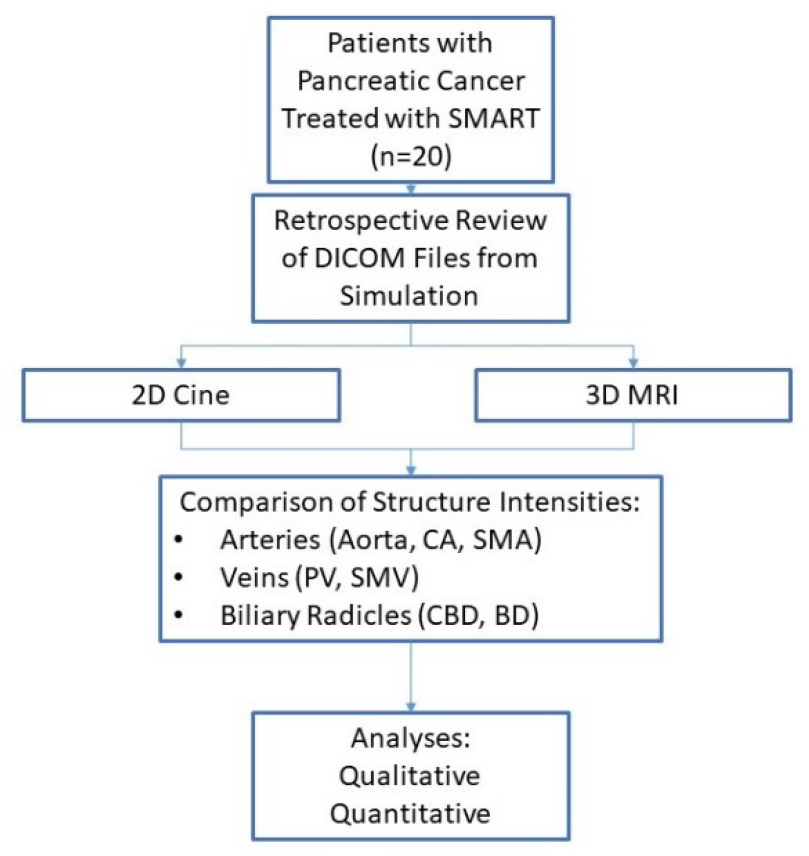
Schema Schema of our methodology for reviewing and comparing the contrast profiles for structures on 2D cine and 3D MRI scans.

### Qualitative Description

We qualitatively identified the relative intensity of arteries (specifically the aorta, celiac axis, and superior mesenteric artery (SMA)), veins (portal vein and superior mesenteric vein (SMV)), and biliary radicles (common bile duct (CBD) and pancreatic duct (PD)).

The relative intensity of each structure was compared between sequences. Any structure that was bright was labeled hyperintense; hyperintensity was subdivided into starkly hyperintense and mildly hyperintense. Any structure that was dark was labeled hypointense. Any structure with similar intensity to surrounding structures or tissues was labeled isointense.

### Quantitative Verification

A custom Python script was used to read the raw image data from MRI DICOM files using the open source PyDicom module (citation: https://zenodo.org/record/5543955#.Y0nnUXbMLHI). Prior to intensity distribution comparison, the 3D MRI volume with in plane resolution of 1.5 × 1.5 mm was resampled to match the in plane resolution of the cine scan (2.4 × 2.4 mm), and the 2D slice closest to the axial location as the cine scan was extracted from each 3D volume. Additionally, a single frame in the deep breath hold position was sampled for comparison to the 3D scan. Pixel intensities values across all images were concatenated to form the two samples: MRI setup scan and MRI cine scan intensities. These samples were then compared using two sample Kolmogorov-Smirnov test available from SciPy (https://scipy.org/citing-scipy/, <u>scipy.stats.ks_2samp — SciPy v1.7.1 Manual</u>). Additionally, the intensity distributions were visualized using a histogram plot generated using Matplotlib (https://matplotlib.org/stable/api/_as_gen/matplotlib.pyplot.hist.html).

## Results

### Case 1

Our first example involves a female patient in her mid 60s who initially presented with weight loss and jaundice. Workup led to diagnosis of a T4N0M0, Stage III (AJCC 8^th^ edition) unresectable adenocarcinoma of the pancreatic head. She underwent 7 cycles of neoadjuvant chemotherapy (4 cycles of FOLFIRINOX, switched to 3 cycles of gemcitabine/paclitaxel due to inadequate response). Restaging workup demonstrated a decreased CA-19-9 and a partial response to chemotherapy on MRI. She then underwent preoperative SMART to the pancreatic head tumor in 50 Gray in 5 fractions. She was slated for pancreaticoduodenectomy which was aborted due to inability to separate the tumor from the common hepatic artery or the replaced right hepatic artery originating from the superior mesenteric artery. The replaced right hepatic artery is indeed visualized in Figure 2.

**Figure 2.**
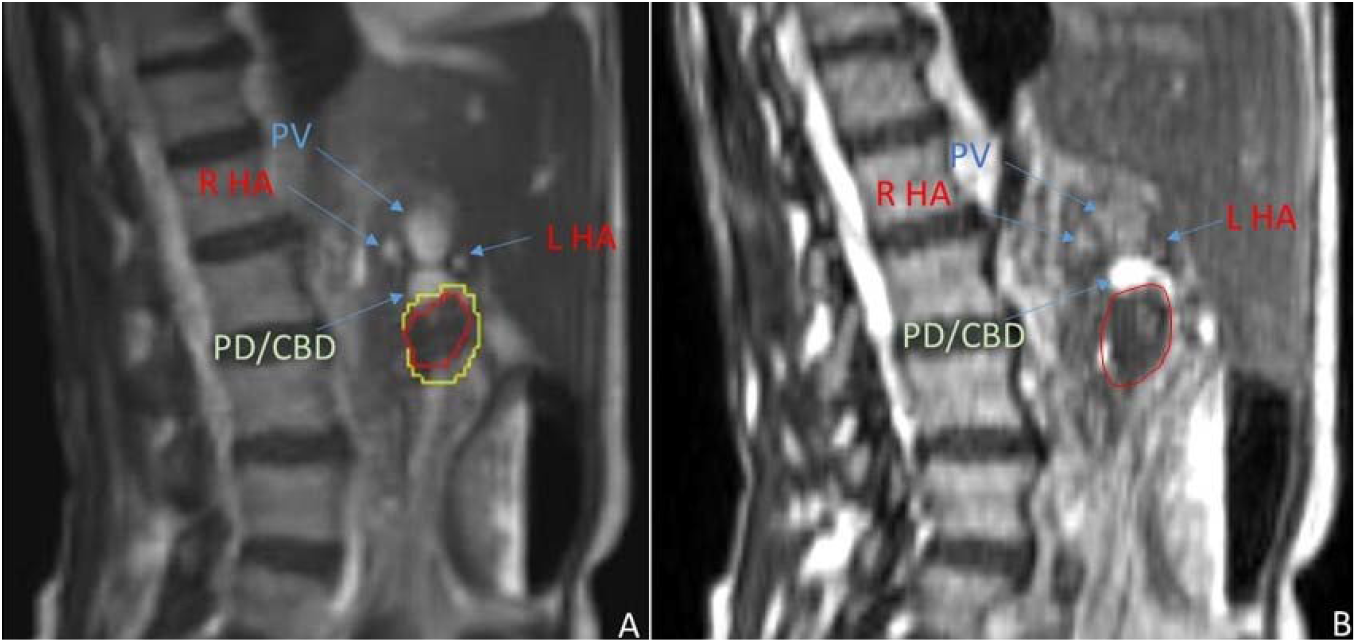
Case 1 Images Comparison of sagittal slices of the 2D cine (panel A) to the corresponding 3D MRI (panel B) for Case 1.

Figure 2 illustrates the qualitative differences in contrast profiles between the 2D cine (panel A) and the 3D MRI (panel B) observed in her case. In both panels, the tumor is seen contoured in red. Additionally, the yellow contour in panel A represents an expansion boundary used for MRgRT beam gating. Superior to the tumor, in both panels, the confluence of the pancreatic duct and the common bile duct is (PD/CBD; light green label) observed as hyperintense; however, in panel B, it is seen as starkly hyperintense. Immediately superior to the PD/CDB confluence, the portal vein (PV; blue label) is seen hyperintense on the 2D cine yet it appears isointense on the 3D MRI. Posterior and anterior to the PV, respectively, the right and left branches of the hepatic artery (R HA, L HA, red) are visualized with similar intensity on both scans.

### Case 2

Our next case is a male in his early 70s who initially presented with dyspepsia, abdominal pain, and weight loss. Workup led to diagnosis of a T4N0M0, Stage III (AJCC 8^th^ edition) borderline resectable ∼3 cm adenocarcinoma of the pancreatic head with involvement of the superior mesenteric artery. He underwent 8 cycles of neoadjuvant mFOLFIRINOX. Restaging workup demonstrated good response via CA-19-9 and CT scan of the abdomen. He subsequently completed preoperative SMART to the pancreatic head tumor in 50 Gray in 5 fractions. He successfully underwent pancreaticoduodenectomy with pathology demonstrating residual ductal adenocarcinoma of the pancreatic head and metastases in 3 of 12 lymph nodes examined, ypT2N1.

His anatomy can be visualized in Figure 3. In both panels, the tumor is seen contoured in red. As in case 1, the yellow contour in panel A represents an expansion boundary used for MRgRT beam gating. Superior to the tumor, in both panels, the superior mesenteric vein is (SMV; blue label) observed as hyperintense; however, in panel B, it is seen as isointense. Immediately anterior to the SMV, the pancreatic duct (PD; light green label) is seen as hyperintense on the 2D cine, yet, it appears starkly hyperintense on the 3D MRI. Superior to the SMV, the common hepatic artery (C HA; red) is visualized to be hyperintense in both scans. Inferior to the tumor, the duodenum can be visualized in both scans (D; orange), and, inferior to the D, the Aorta (AO; red) can be visualized as hyperintense in both scans.

**Figure 3.**
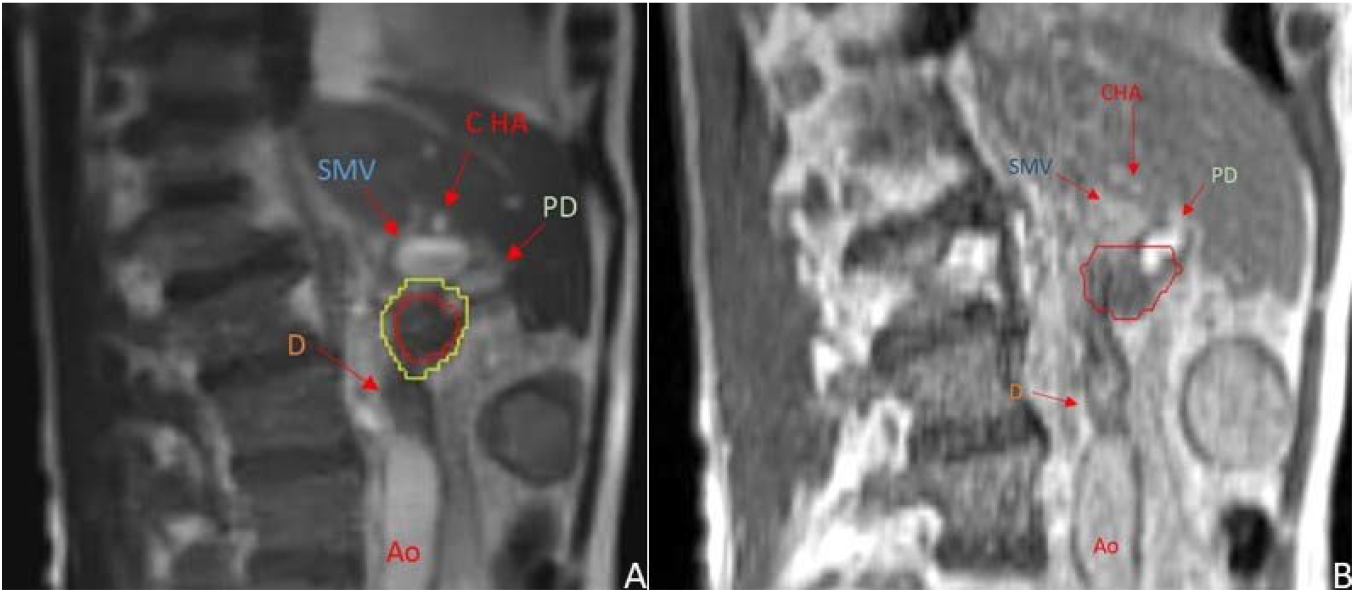
Case 2 Images Comparison of sagittal slices of the 2D cine (panel A) to the corresponding 3D MRI (panel B) for Case 2.

### Case 3

Our final case is a male in his late 60s who initially presented with abdominal pain and jaundice. Workup led to diagnosis of a T2N0M0, Stage II (AJCC 8^th^ edition) borderline resectable 3.5 cm adenocarcinoma of the pancreatic head. He underwent biliary stenting followed by 7 cycles of neoadjuvant mFOLFIRINOX. Restaging workup demonstrated good response via CA-19-9 and CT of the abdomen. He subsequently completed preoperative SMART to the pancreatic head tumor in 50 Gray in 5 fractions. He underwent pancreaticoduodenectomy successfully with pathology demonstrating residual ductal adenocarcinoma of the pancreatic head, ypT2N0.

His anatomy can be visualized in Figure 4. In both panels, the tumor is seen contoured in red. As in our prior cases, the yellow contour in panel A represents an expansion boundary used for MRgRT beam gating. Superior to the tumor, in both panels, the common bile duct is (BD; light green label) observed as hyperintense; however, in panel B, it is seen as starkly hyperintense. Immediately superior and posterior to the BD, the portal vein (PV; blue label) is seen hyperintense on the 2D cine yet it appears isointense on the 3D MRI. Superior to the PV, the common hepatic artery (HA; red) is visualized to be mildly hyperintense in both scans. Anterior to the tumor, the duodenum is visualized in both scans (D; orange). Posterior to the tumor, the inferior vena cava (IVC; blue) is visualized as hyperintense on the 2D cine yet isointense on the 3D MRI.

**Figure 4.**
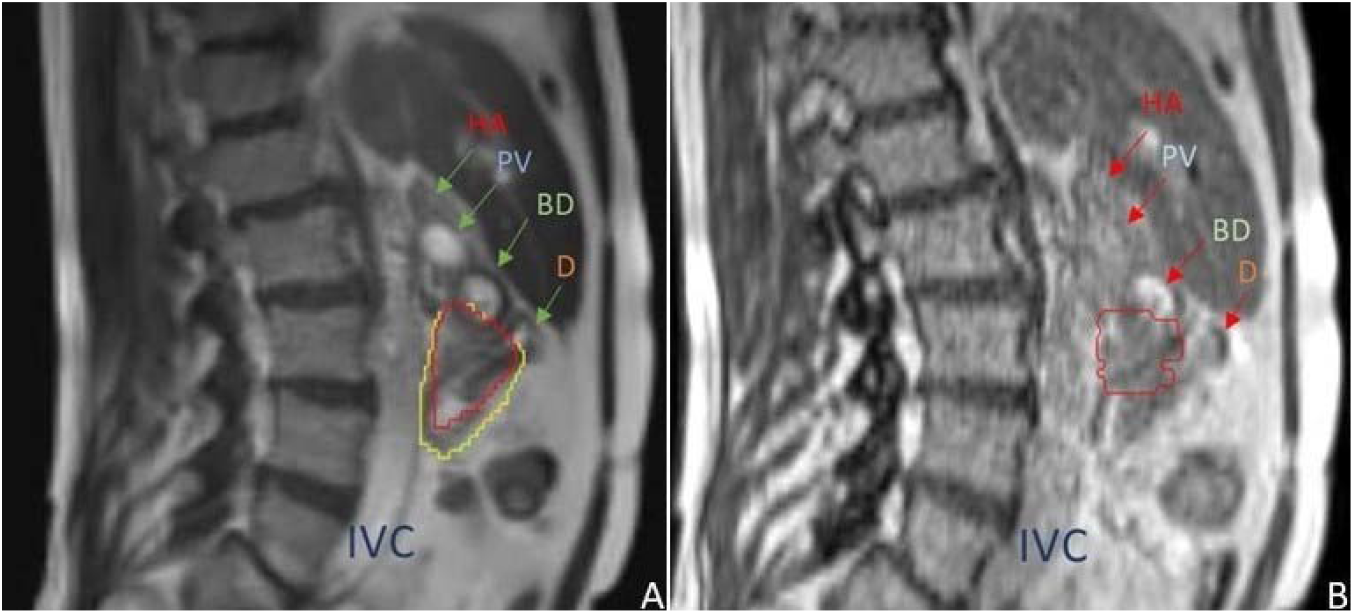
Case 3 Images Comparison of sagittal slices of the 2D cine (panel A) to the corresponding 3D MRI (panel B) for Case 3.

### Qualitative Findings

In all 20 patients reviewed, arteries (aorta, celiac, SMA, HA) typically appeared with mild hyperintensity resulting in similar contrast profiles in both scans. However, veins (PV, SMV) appeared consistently hyperintense on 2D cine and consistently isointense on 3D MRI. Biliary structures (CBD, PD) appeared consistently hyperintense on both scans, only mildly hyperintense on 2D cine but starkly hyperintense on 3D MRI. These findings are summarized in Table 1.

**Table 1.** Results Qualitative Results of Clinically Observed Intensity of Arteries, Veins, and Biliary Radicles on both 2D Cine and 3D MRI sequences.

| STRUCTURE REVIEWED | 2D CINE | 3D MRI |
| --- | --- | --- |
| Arteries (aorta, celiac, SMA, HA) | Mildly Hyperintense | Mildly Hyperintense |
| Veins (PV, SMV) | Hyperintense | Isointense |
| Biliary Radicles (CBD, PD) | Mildly Hyperintense | Starkly Hyperintense |

### Quantitative Verification

The pixel intensity distributions extracted from 2D cine and 3D MRI images were confirmed to differ significantly (two sample Kolmogorov-Smirnov test; test statistic=0.40; p<0.001) in Figure 5.

**Figure 5.**
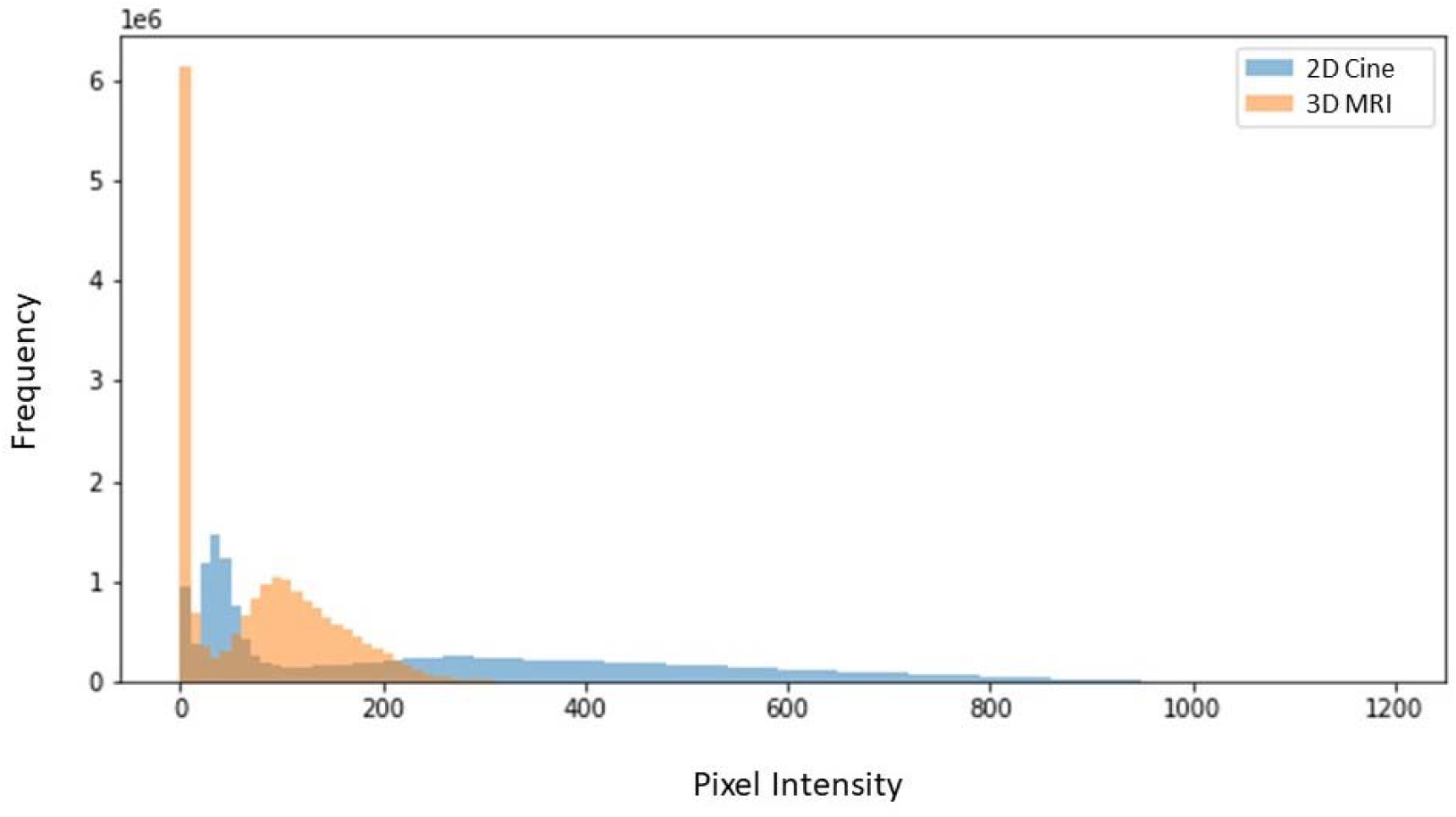
Pixel Intensity Histogram Comparison of Pixel intensity distributions for 2D cine and 3D MRI images.

## Discussion

Our findings qualitatively confirm our hypothesis that arteries, veins, and biliary radicles do indeed appear with different intensities on 2D cine versus the 3D MRI during SMART. Additionally, Figure 5 demonstrates the disparity in pixel intensity between the two data sets. We hypothesize that this difference in intensity may have to do with the biophysical characteristics of the fluid content including velocity of fluid and/or oxygen content of the conduits, particularly in arteries.

Understanding these significant variations of image intensity between the immediate pre-treatment 2D cine and the initial treatment planning 3D MRI can guide radiation oncologists in choosing optimal tracking targets. For example, if a tracking structure is selected due to its relative intensity to the background based on the 3D MRI this may, in fact, not be consistent with the relative intensity to background on 2D cine and lead to tracking target failure which could lead to improper/inefficient gating, or need for contouring an alternative tracking target. Any of these scenarios could potentially cause confusion or uncertainty of the treatment team at the time of radiation delivery. Furthermore, all of these situations could lead to increased time for delivery and increased time on table in an enclosed MR-linac.

The ViewRay system is slated to upgrade to 3D target tracking. Although this would not affect the contrast profiles of the 2D cine or the 3D MRI, it would allow for more information to confirm anatomical identity of structures due to the ability to track with a 3D cine. Lastly, our group is looking at exploring alternative tracking algorithms using artificial intelligence and deep learning which could improve the ViewRay user experience.

## Conclusions

Understanding variations of image intensity between the different MRI sequences used in SMART, or MRgRT at large, is valuable to radiation oncologists and may lead to improved target tracking.

## Data Availability

All data produced in the present work are contained in the manuscript.

## Acknowledgements

Charles R. Thomas, Jr, MD

## References

1. Siegel RL, Miller KD, Fuchs HE, Jemal A. Cancer statistics, 2022. CA Cancer J Clin. 2022;72(1):7–33. doi:10.3322/caac.21708

2. Versteijne E, van Dam JL, Suker M, et al. Neoadjuvant Chemoradiotherapy Versus Upfront Surgery for Resectable and Borderline Resectable Pancreatic Cancer: Long-Term Results of the Dutch Randomized PREOPANC Trial. J Clin Oncol. 2022;40(11):1220–1230. doi:10.1200/JCO.21.02233

3. Hammel P, Huguet F, van Laethem JL, et al. Effect of Chemoradiotherapy vs Chemotherapy on Survival in Patients With Locally Advanced Pancreatic Cancer Controlled After 4 Months of Gemcitabine With or Without Erlotinib: The LAP07 Randomized Clinical Trial. JAMA. 2016;315(17):1844–1853. doi:10.1001/jama.2016.4324

4. Zhong J, Patel K, Switchenko J, et al. Outcomes for patients with locally advanced pancreatic adenocarcinoma treated with stereotactic body radiation therapy versus conventionally fractionated radiation: SBRT Versus Conventional RT in LAPC. Cancer. 2017;123(18):3486–3493. doi:10.1002/cncr.30706

5. Rudra S, Jiang N, Rosenberg SA, et al. Using adaptive magnetic resonance image-guided radiation therapy for treatment of inoperable pancreatic cancer. Cancer Med. 2019;8(5):2123–2132. doi:10.1002/cam4.2100

6. Petrelli F, Comito T, Ghidini A, Torri V, Scorsetti M, Barni S. Stereotactic Body Radiation Therapy for Locally Advanced Pancreatic Cancer: A Systematic Review and Pooled Analysis of 19 Trials. Int J Radiat Oncol. 2017;97(2):313–322. doi:10.1016/j.ijrobp.2016.10.030

7. Mazzarotto R, Simoni N, Guariglia S, et al. Dosimetric Feasibility Study of Dose Escalated Stereotactic Body Radiation Therapy (SBRT) in Locally Advanced Pancreatic Cancer (LAPC) Patients: It Is Time to Raise the Bar. Front Oncol. 2020;10:600940. doi:10.3389/fonc.2020.600940

8. Katz MHG, Shi Q, Meyers J, et al. Efficacy of Preoperative mFOLFIRINOX vs mFOLFIRINOX Plus Hypofractionated Radiotherapy for Borderline Resectable Adenocarcinoma of the Pancreas: The A021501 Phase 2 Randomized Clinical Trial. JAMA Oncol. 2022;8(9):1263–1270. doi:10.1001/jamaoncol.2022.2319

9. Janssen QP, van Dam JL, Prakash LR, et al. Neoadjuvant Radiotherapy After (m)FOLFIRINOX for Borderline Resectable Pancreatic Adenocarcinoma: A TAPS Consortium Study. J Natl Compr Canc Netw. 2022;20(7):783–791.e1. doi:10.6004/jnccn.2022.7008

10. Chuong MD, Bryant J, Mittauer KE, et al. Ablative 5-Fraction Stereotactic Magnetic Resonance– Guided Radiation Therapy With On-Table Adaptive Replanning and Elective Nodal Irradiation for Inoperable Pancreas Cancer. Pract Radiat Oncol. 2021;11(2):134–147. doi:10.1016/j.prro.2020.09.005

11. Parikh PJ, Lee P, Low D, et al. Stereotactic MR-Guided On-Table Adaptive Radiation Therapy (SMART) for Patients with Borderline or Locally Advanced Pancreatic Cancer: Primary Endpoint Outcomes of a Prospective Phase II Multi-Center International Trial. Int J Radiat Oncol. 2022;114(5):1062–1063. doi:10.1016/j.ijrobp.2022.09.010

12. Herman JM, Chang DT, Goodman KA, et al. Phase 2 multi-institutional trial evaluating gemcitabine and stereotactic body radiotherapy for patients with locally advanced unresectable pancreatic adenocarcinoma. Cancer. 2015;121(7):1128–1137. doi:10.1002/cncr.29161

13. Reyngold M, Parikh P, Crane CH. Ablative radiation therapy for locally advanced pancreatic cancer: techniques and results. Radiat Oncol. 2019;14(1):95. doi:10.1186/s13014-019-1309-x

14. Zur Y, Bosak E, Kaplan N. A new diffusion SSFP imaging technique. Magn Reson Med. 1997;37(5):716–722. doi:10.1002/mrm.1910370513

15. Zur Y, Wood ML, Neuringer LJ. Motion-insensitive, steady-state free precession imaging. Magn Reson Med. 1990 Dec;16(3):444–59.

